# ADMINISTRATION OF LEVODOPA IN A PARKINSON’S PATIENT USING A NOVEL ORAL AND MAXILLOFACIAL ROUTE – A FIRST IN HUMAN STUDY

**DOI:** 10.1101/2023.07.13.23292411

**Authors:** Suresh Thirunavukarasu, Balasubramanian Bala Venkata Ramanan, Sathya Krishnan Suresh, Vincent Jayakumar Antonisamy, Devi Varadharaj, Paranjothi Shanmugam, Kavita Verma, Canmany Elumalai, Gladson Selvakumar, Ahila Elumalai, Lydia Prabahar, Hridwik Adiyeri Janardhanan, Anoop UR

## Abstract

Parkinson’s disease is characterized by loss of nigrostriatal dopaminergic neurons in the brain. Dopamine cannot be administered systemically as a treatment because dopamine does not cross the blood-brain-barrier. Oral levodopa is the gold standard till date. Currently, in patients who show poor response to oral levodopa, the drug can be delivered through alternate routes like inhalation and continuously by intestinal and subcutaneous routes. In this report, a novel oral and maxillofacial route was used for the first time in the world to administer levodopa in a Parkinson’s patient and its efficacy was compared with the oral route of administration.

## INTRODUCTION

Parkinson’s disease is the second most common neurodegenerative disorder^1^.Oral levodopa has been the gold standard in the treatment of Parkinson’s disease^2,3^. Oral levodopa is decarboxylated in the gut and only about 30% of levodopa crosses the intestinal mucosa. The absorbed levodopa is further metabolized by the liver, the kidney and by the brain capillary endothelium. The effectiveness of levodopa depends on its metabolism to dopamine in the brain. But only about 1% of oral levodopa crosses the blood brain barrier^4^.

Levodopa provides good symptom relief in the initial years. But as the disease progresses, the effect of levodopa decreases, and motor complications occur. Such patients require more levodopa at shorter intervals. As the therapeutic window also narrows with disease progression, the higher daily levodopa dose results in levodopa induced dyskinesia because of the higher levodopa concentrations, the higher levodopa peaks and pulsatile dopaminergic stimulation^5^. The higher levels of levodopa metabolites in the periphery causes adverse effects like orthostatic hypotension, nausea and vomiting^4^.

Addition of peripheral enzyme inhibitors increase the plasma half-life and decrease the dose of levodopa^5^.Alternative approved levodopa delivery options for those patients who show poor response to oral levodopa are intestinal gel pump therapy and inhalable levodopa ^6,7^. Sub-cutaneous levodopa infusion therapy is yet to be approved. These therapies have limitations and there is still a need for a simple and painless technique to deliver low dose anti-Parkinson’s drugs across the blood brain barrier without systemic complications.

Parkinson’s patients have a higher incidence of dental caries, periodontal diseases and loss of teeth^8^. Extraction of non-vital teeth or teeth with dental caries involving the pulp can be avoided by doing root canal treatment.

In this report, levodopa administered through a novel oral and maxillofacial route in a consenting Parkinson’s disease patient is compared with levodopa/carbidopa administered through the oral route.

## METHOD

A Parkinson’s patient attending the Movement Disorders Clinic at Department of Neurology, Indira Gandhi Government General Hospital and Post Graduate Institute, Puducherry, India was selected for the study. The patient also had a non-vital upper posterior tooth that required root canal treatment.

### Pre-study Examination

On clinical examination, the vitals were stable, blood pressure was 120/ 80 mm of Hg with no postural fall in blood pressure, pulse was70/minute regular in volume and rhythm., Higher mental function was normal with MMSE of 28/30. Cranial nerve examination was normal. Spino-motor system examination showed normal. Bulk and power were 5/5 for all four limbs. The patient had cogwheel rigidity of upper and lower limbs which was more distally. Deep tendon reflexes were normally elicitable and plantar was flexor bilaterally. Extrapyramidal system examination showed mask like facies with decreased blink rate. Rest and postural tremor were more on the left side when compared to the right side and the patient also had short-stepping gait with festination, enbloc turning and pull test was positive. The patient satisfied the UK brain bank criteria for Parkinson’s disease and was started on Syndopa initially at 110 mg twice a day and increased to thrice a day after one year. Neuroimaging including CT scan of the brain was normal and the patient had good response to Syndopa.

Oral examination revealed multiple root stumps and carious teeth. The upper right second premolar was non-vital. Radiographic examination showed the apex of the non-vital tooth in close proximity to the floor of the maxillary sinus.

### Study Procedure

Root canal treatment was initiated and the pulp cavity was debrided, cleaned and enlarged. A digital scan of the treated tooth was made. A 3D printed oral and maxillofacial drug delivery system that could be attached to the tooth was designed to deliver the drug through the tooth.

The patient reported in the morning for two consecutive days without taking the morning dose of levodopa/carbidopa and without taking breakfast. A wash out period of minimum 7.5 hours was maintained between the last dose of oral levodopa/carbidopa and administration of the drug at the hospital.

On each day, the pre-dose blood sample was taken and the pre-dose UPDRS (Unified Parkinson’s Disease Rating Scale, part 3, the motor component was assessed). On the first day, the patient was administered the morning dose of levodopa/carbidopa tablet and the patient had breakfast one hour after the drug administration. On the second day, the patient was administered 1/20^th^ of the oral dose of levodopa through the oral and maxillofacial route using an oral and maxillofacial drug delivery system connected to the root canal treated tooth. The drug delivery system was removed after drug delivery. The patient had breakfast one hour after the drug administration. The breakfast menu was the same on both the days.

Blood samples were taken and UPDRS scores were assessed at 30 minutes, 60 minutes, 90 minutes and 120 minutes after drug administration. The blood samples were centrifuged and the plasma was separated and stored in dry ice at -80C. The blood samples were then subjected to HPLC.

## RESULT

Blood values of levodopa after oral tablet peaked at 30 minutes and fell down to negligible levels by 60 minutes. The oral and maxillofacial route showed a higher peak at 35 minutes and decreased from 60 minutes to 120 minutes. The UPDRS score was higher for the oral route and lower for the oral and maxillofacial route. (Figure:1).

**Figure:1.**
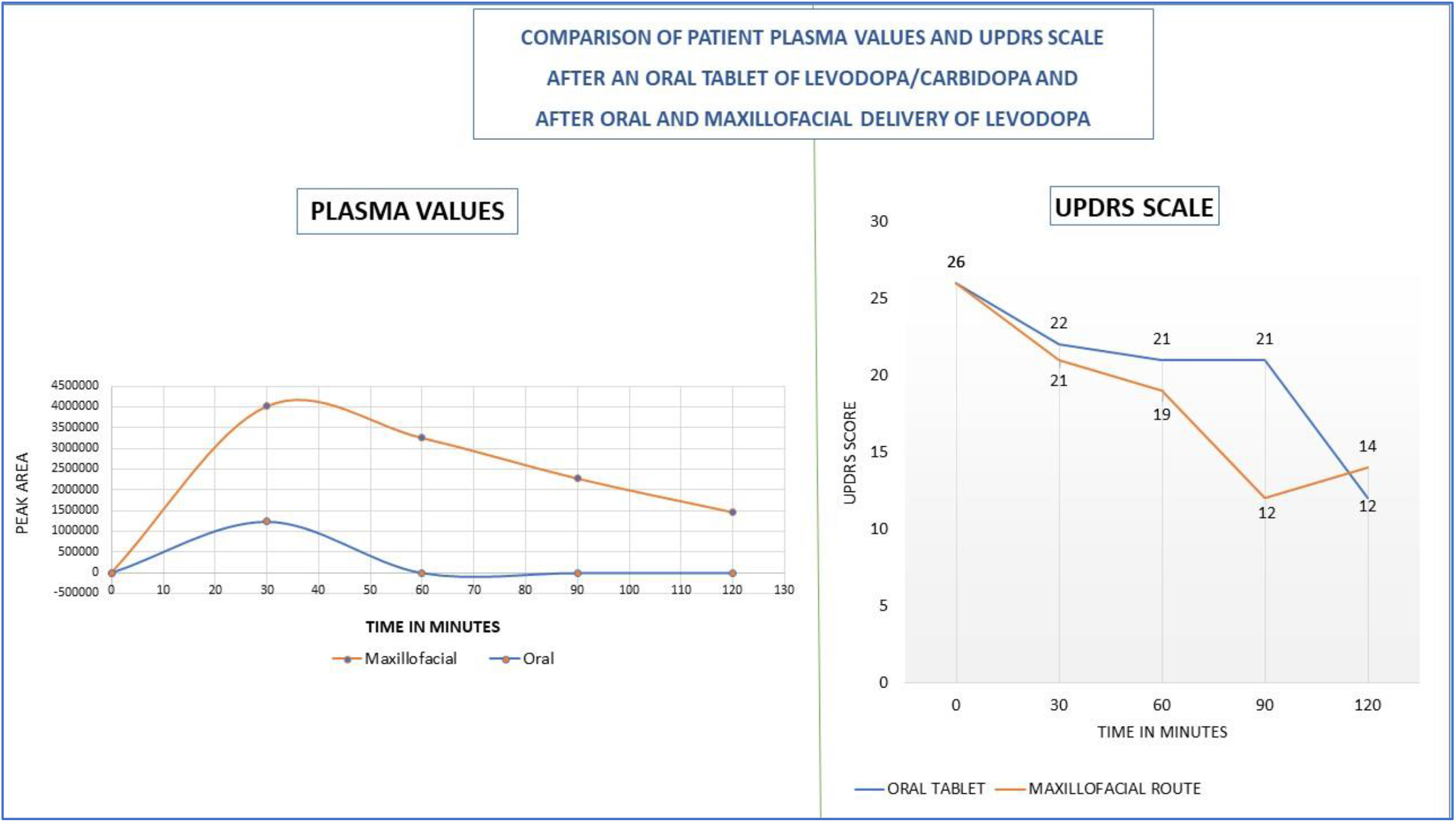

The pharmacokinetic analysis was done using the linear trapezoidal method and the area under the concentration time curve (AUC) was calculated. The relative bioavailability of the oral and maxillofacial route was consistently higher in the plasma level starting at the 5^th^ minute and was maintained till 120^th^ minute as shown by (Figure :2).

**Figure:2.**
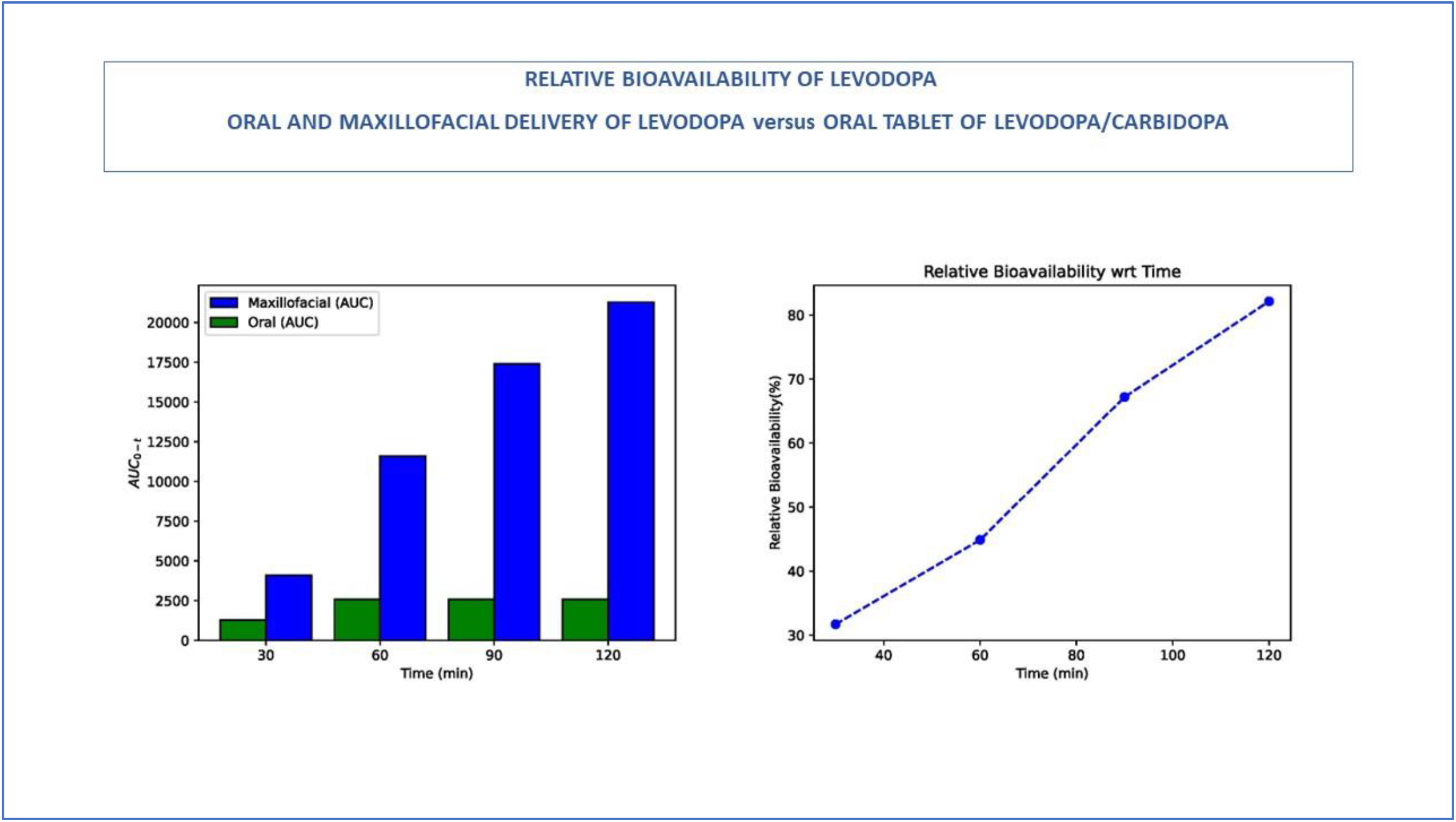

The motor component of UPDRS showed a one-point difference at 30^th^ minute favoring the oral and maxillofacial route. At 60^th^ minute it improved by two more points. When compared with the oral route, the best scores in favor of the oral and maxillofacial route were noted at the 90^th^ minute. A relatively high plasma value at 30^th^ minute following oral and maxillofacial administration correlated only with mild clinical improvement in UPDRS, motor component. But subsequently the patient had good clinical improvement till 90^th^ minute as noted in UPDRS scores, which also corresponded with the high plasma values following oral and maxillofacial administration.

This result may suggest that the oral and maxillofacial route may be useful for frequent low dosing to prevent off phenomenon as well as dyskinesia. The efficacy of the oral and maxillofacial route as on-demand treatment for early morning OFF, delayed ON and no ON requires further evaluation and assessment because the clinical improvement at 30^th^ minute was not high in the oral and maxillofacial route when compared with the oral route despite having a high plasma value. As this study is limited to one patient, a clearer understanding will emerge when frequent doses of levodopa are administered in more patients to measure the exact outcome.

## DISCUSSION

James Parkinson’s original description of the “Shaking Palsy” was first published in 1817. Levodopa was introduced 140 years later^9^. It provides only symptomatic relief. Even after more than 200 years, there is still no curative treatment for Parkinson’s.

Parkinson’s disease is a progressive, neurodegenerative disorder caused by the loss of dopaminergic neurons from the substantia nigra pars compacta in the brain^4^. Parkinson’s disease is now considered a recognizable clinical syndrome and is defined by the presence of bradykinesia combined with either rest tremor, rigidity, or both. Currently, no therapy slows down the progression and there is a need for personalized management^10^.An integrated approach includes drug treatment alongside device-aided therapies, multidisciplinary care and patient empowerment^11^.

Levodopa is still the gold standard in the treatment of Parkinson’s disease. Several factors affect the availability of levodopa. Orally given levodopa is absorbed in the proximal one-third of the small intestine.30%-100% of PD patients suffer from impaired gastric motility. Delayed delivery of levodopa to the intestinal absorption sites results in increased pre-systemic peripheral decarboxylation and reduced levodopa absorption. It contributes to ‘delayed on’, ‘dose failure’ and to medication overload. The fluctuating levodopa levels result in increased motor complications^12^.Dietary amino acids and gut bacterial interference with levodopa treatment can also reduce the absorption of levodopa in the small intestine.^13^.

Despite its relatively short half-life, levodopa is effective in the early stages of the disease probably due to the preserved capacity of the presynaptic nerve terminals to store dopamine. The effect of levodopa progressively gets shorter with continued loss of neurons in the substantia nigra. This can initially be improved by increasing the frequency of levodopa doses. With further progression, motor fluctuations, ‘‘on/ off’’ phenomena, and dyskinesias appear which may not respond to the increased frequency of levodopa doses. The fluctuating plasma levels of levodopa result in akinesia at subtherapeutic levels and dyskinesias at peak levels. With further progression, dyskinesias become more complex with practically no response to levodopa^14^.

Modifications to levodopa formulations, use of complimentary agents that improve levodopa bioavailability and device-assisted approaches can be used to treat OFF periods. Patients will ultimately require device assisted therapies to manage motor fluctuations.^6.^

Levodopa has a short half-life of about 50 minutes. Addition of peripheral enzyme inhibitors increase the plasma half-life to about 90 minutes and decrease the required dose of levodopa. Combining levodopa with carbidopa or benserazide decreases the oral dose by 80% and increases the amount of levodopa available to the brain. ^13^

Exogenous levodopa having a short half-life causes intermittent release of dopamine from the presynaptic neurons resulting in non-physiological, pulsatile stimulation of the postsynaptic receptors. The pulsatile stimulation produces changes in the postsynaptic receptors resulting in motor complications ^14^. Clinical evidence suggests that continuous delivery of levodopa can reduce the motor complications. A study reported that continuous oral delivery of levodopa/carbidopa was associated with less plasma variability and reduced off time in comparison to standard intermittent oral levodopa/carbidopa therapy^15.^ Controlled release oral formulations show sustained plasma levels than standard levodopa/ carbidopa. The drawbacks include lower bioavailability, longer time to peak, delayed onset of clinical response and need for a higher daily dose in spite of a lower frequency of dosing.^2,3^

Newer routes of levodopa administration are also used to provide continuous delivery. Intraduodenal route bypasses the stomach and delivers levodopa directly to its absorption sites in the small intestine resulting in a stable plasma concentration when compared with intermittent oral dosing. It reduces the OFF time as well as the disabling dyskinesias. The common adverse effects were related either to the surgical procedure or the device ^2,3^. Levodopa/carbidopa intestinal gel infusion resulted in faster absorption than oral tablets and reduced intra-subject variability.^16^

Poor levodopa solubility is a challenge in the development of subcutaneous formulations.^2,3^An open label study has stated continuous subcutaneous levodopa /carbidopa as a feasible route ^17^. A comparison between continuous subcutaneous infusion, intravenous infusion using a continuously buffered acidic levodopa/carbidopa and intestinal levodopa/carbidopa gel infusion yielded similar levodopa levels at 30 minutes. ^18^

Recently, an inhalable formulation of levodopa has been approved for intermittent treatment of OFF periods in patients treated with levodopa/carbidopa. Oral inhalation bypasses the gut and levodopa can be delivered into the lungs for rapid absorption into the systemic circulation. It is not recommended in patients with asthma or other chronic lung diseases. Cough was the most common adverse event. Improvement in UPDRS-III score was noted at 10 minutes and was sustained till 60 minutes^7^. Orally inhaled levodopa without a dopa decarboxylase inhibitor was compared with oral levodopa/carbidopa. Inhaled levodopa was absorbed faster than oral levodopa with a bioavailability of 53%^19.^ The faster absorption helped in tiding over the off periods which was demonstrated in two studies.^20,21.^

In this report, we present a novel oral and maxillofacial route for delivering levodopa. In vivo animal studies have shown that drugs can be successfully delivered into the brain through this route by using the paravascular, vascular, neural, lymphatic and glymphatic pathways.^22, 23, 24.^

The frequency of untreated caries in patients with Parkinson’s is reported as high at Hoehn and Yahr stage II and above.^25^ In this study, levodopa was administered in the Parkinson’s patient through an upper root canal treated tooth using a novel 3D printed oral and maxillofacial drug delivery system that was temporarily connected to the tooth. The novel route showed significant advantages over the oral route. A comparison of different routes is shown in (Table:1).

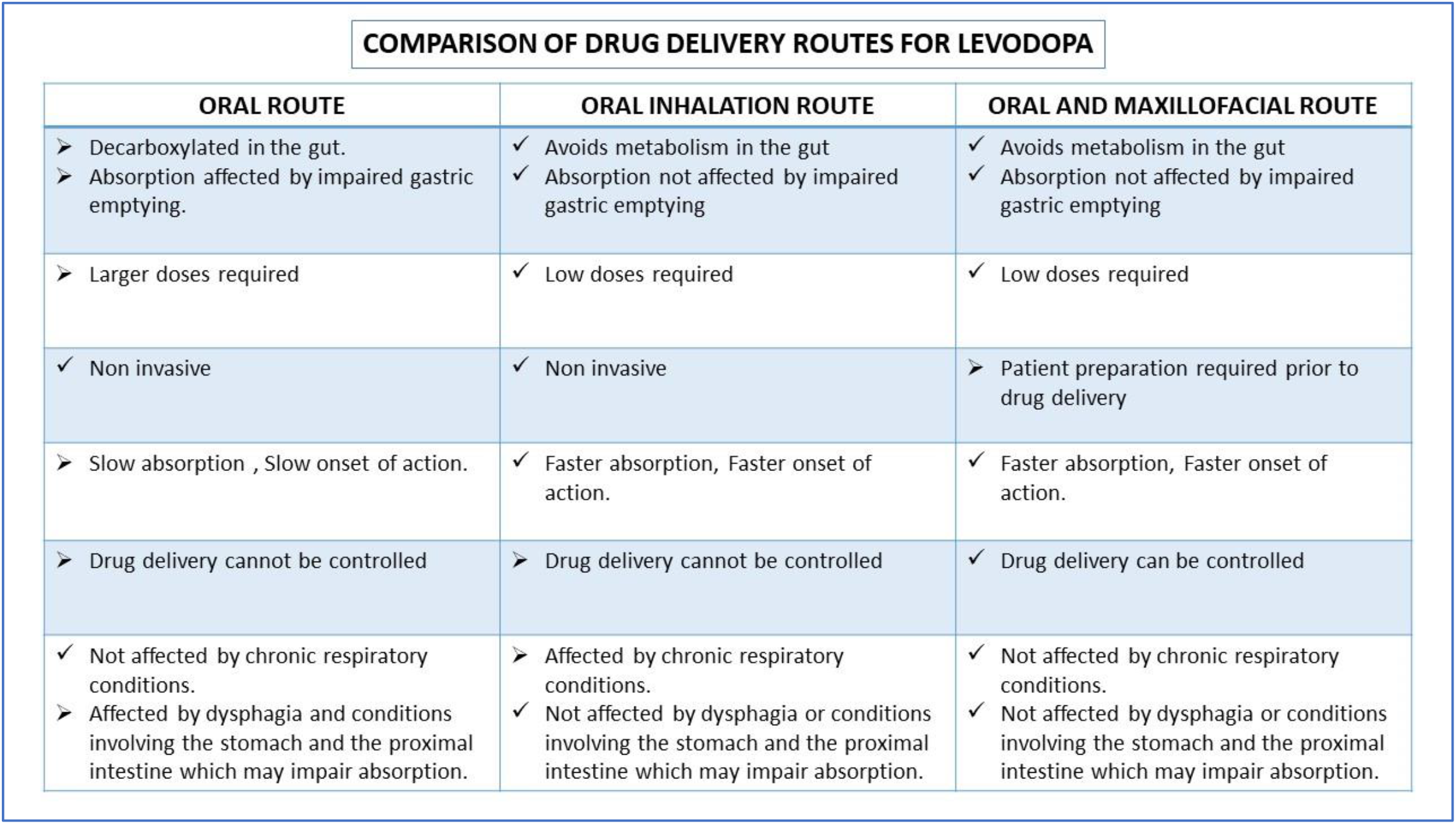

## CONCLUSION

Low doses of levodopa can be administered painlessly through this novel oral and maxillofacial route by bypassing metabolism in the gut, first pass metabolism in the liver and metabolism in the brain capillary endothelium.

This is a single case study showing good absorption of levodopa resulting in a steady state in plasma with corresponding good UPDRS scores. Apart from being a novel way of administering levodopa, we hope the route can help patients with dementia, patients who are bedridden requiring help for activities of daily life, patients in the ICU set up with Ryle’s tube in situ and in patients with dyskinesias where frequent low doses will be required to attain a steady state in the plasma. Though further studies are required, the oral and maxillofacial route of administration can also be an on-demand treatment option for early morning offs, end-of-dose wearing offs, delayed on, suboptimal on and dose failure.

## Statements And Declarations

### Ethics approval

The research protocol was approved by the Institutional Ethics Committee, Indira Gandhi Government General Hospital and Post Graduate Institute Ref No: IGGGH&PGI/Estt/E15/2023/149 dated 31/03/2023.

The study has been registered prospectively with ICMR Clinical Trial Registry - India as CTRI/2023/04/051539.

The human participant has given written informed consent.

### Consent to participate

Consent Taken

### Consent to publish

Consent Taken

### Data Availability

Clinical data produced in the present study are available upon reasonable request to the corresponding author.

### Author’s contribution

Author **1** is the Principal Investigator of the study. **1, 7 & 13** contributed to the conception, acquisition, analysis, interpretation of data, drafting, reviewing and final approval. **3** contributed to the acquisition, analysis, interpretation of data, drafting, reviewing and final approval. **2**,**4**,**5**,**6** contributed to the analysis of medical data, reviewing and approval. **8**,**9**,**10**,**11**,**12** contributed in acquisition of dental data, reviewing and approval.

## Acknowledgment

We acknowledge Dr.D.Saravanan, Phd, co-ordinator, National College Instrumentation Facility, Tiruchirapalli, India and Mrs.S.Kavitha, M Pharm, HPLC analyst, National College Instrumentation Facility, Tiruchirapalli, India for their invaluable support in completing this study. We acknowledge Mr Seenukumar.S and Mr.Murugamani.K for their support.

## REFERENCES

1. Aarsland, D., Batzu, L., Halliday, G.M., Geurtsen, G.J., Ballard, C., Ray Chaudhuri, K. and Weintraub, D., 2021. Parkinson disease-associated cognitive impairment. Nature Reviews Disease Primers, 7(1), p.47

2. Freitas ME, Ruiz-Lopez M, Fox SH. Novel Levodopa Formulations for Parkinson’s Disease. CNS Drugs. 2016 Nov;30(11):1079–1095. doi: 10.1007/s40263-016-0386-8. PMID: 27743318.

3. Pilleri M, Antonini A. Novel levodopa formulations in the treatment of Parkinson’s disease. Expert Rev Neurother. 2014 Feb;14(2):143–9. doi: 10.1586/14737175.2014.877840. Epub 2014 Jan 13. PMID: 24428803.

4. Hoon, M., Petzer, J.P., Viljoen, F. and Petzer, A., 2017. The Design and Evaluation of an L-Dopa–Lazabemide Prodrug for the Treatment of Parkinson’s Disease. Molecules, 22(12), p.2076.

5. Bandopadhyay R, Mishra N, Rana R, Kaur G, Ghoneim MM, Alshehri S, Mustafa G, Ahmad J, Alhakamy NA, Mishra A. Molecular Mechanisms and Therapeutic Strategies for Levodopa-Induced Dyskinesia in Parkinson’s Disease: A Perspective Through Preclinical and Clinical Evidence. Front Pharmacol. 2022 Apr 7; 13:805388. doi: 10.3389/fphar.2022.805388. PMID: 35462934; PMCID: PMC9021725.

6. Vijiaratnam N, Foltynie T. Therapeutic Strategies to Treat or Prevent Off Episodes in Adults with Parkinson’s Disease. Drugs. 2020 Jun;80(8):775–796. doi: 10.1007/s40265-020-01310-2. PMID: 32382948.

7. Hauser RA, LeWitt PA, Comella CL. On demand therapy for Parkinson’s disease patients: Opportunities and choices. Postgrad Med. 2021 Sep;133(7):721–727. doi: 10.1080/00325481.2021.1936087. Epub 2021 Jun 30. PMID: 34082655.

8. Bollero P, Franco R, Cecchetti F, Miranda M, Barlattani A Jr, Dolci A, Ottria L. Oral health and implant therapy in Parkinson’s patients: review. Oral Implantol (Rome). 2017 Sep 27;10(2):105–111. doi: 10.11138/orl/2017.10.2.105. PMID: 29876035; PMCID: PMC5965075.

9. Palacios-Sánchez, L., Torres Nupan, M. and Botero-Meneses, J.S., 2017. James Parkinson and his essay on “shaking palsy”, two hundred years later.Arquivos de Neuro-Psiquiatria,75, pp.671–672.

10. Bloem BR, Okun MS, Klein C. Parkinson’s disease. Lancet. 2021 Jun 12;397(10291):2284–2303. doi: 10.1016/S0140-6736(21)00218-X. Epub 2021 Apr 10. PMID: 33848468.

11. Beckers M, Bloem BR, Verbeek MM. Mechanisms of peripheral levodopa resistance in Parkinson’s disease. NPJ Parkinsons Dis. 2022 May 11;8(1):56. doi: 10.1038/s41531-022-00321-y. PMID: 35546556; PMCID: PMC9095610.

12. Leta V, Klingelhoefer L, Longardner K, et al. Gastrointestinal barriers to levodopa transport and absorption in Parkinson’s disease. Eur J Neurol.2023; 30:1465–1480. doi:10.1111/ene.15734

13. van Kessel SP, El Aidy S. Contributions of Gut Bacteria and Diet to Drug Pharmacokinetics in the Treatment of Parkinson’s Disease. Front Neurol. 2019 Oct 15; 10:1087. doi: 10.3389/fneur.2019.01087. PMID: 31681153; PMCID: PMC6803777.

14. Thanvi BR, Lo TC. Long term motor complications of levodopa: clinical features, mechanisms, and management strategies. Postgrad Med J. 2004 Aug;80(946):452–8. doi: 10.1136/pgmj.2003.013912. PMID: 15299154; PMCID: PMC1743071.

15. Warren Olanow C, Torti M, Kieburtz K, Leinonen M, Vacca L, Grassini P, Heller A, Heller E, Stocchi F. Continuous versus intermittent oral administration of levodopa in Parkinson’s disease patients with motor fluctuations: A pharmacokinetics, safety, and efficacy study. Mov Disord. 2019 Mar;34(3):425–429. doi: 10.1002/mds.27610. Epub 2019 Jan 17. PMID: 30653246.

16. Othman AA, Dutta S. Population pharmacokinetics of levodopa in subjects with advanced Parkinson’s disease: levodopa-carbidopa intestinal gel infusion vs. oral tablets. Br J Clin Pharmacol. 2014 Jul;78(1):94–105. doi: 10.1111/bcp.12324. PMID: 24433449; PMCID: PMC4168384.

17. Olanow CW, Espay AJ, Stocchi F, Ellenbogen AL, Leinonen M, Adar L, Case RJ, Orenbach SF, Yardeni T, Oren S, Poewe W; 006 study group. Continuous Subcutaneous Levodopa Delivery for Parkinson’s Disease: A Randomized Study. J Parkinsons Dis. 2021;11(1):177–186. doi: 10.3233/JPD-202285. PMID: 33164945; PMCID: PMC7990424.

18. Bergquist F, Ehrnebo M, Nyholm D, Johansson A, Lundin F, Odin P, Svenningsson P, Hansson F, Bring L, Eriksson E, Dizdar N. Pharmacokinetics of Intravenously (DIZ101), Subcutaneously (DIZ102), and Intestinally (LCIG) Infused Levodopa in Advanced Parkinson Disease. Neurology. 2022 Jun 15;99(10): e965–76. doi: 10.1212/WNL.0000000000200804. Epub ahead of print. PMID: 35705502; PMCID: PMC9519246.

19. Luinstra M, Rutgers W, van Laar T, Grasmeijer F, Begeman A, Isufi V, Steenhuis L, Hagedoorn P, de Boer A, Frijlink HW. Pharmacokinetics and tolerability of inhaled levodopa from a new dry-powder inhaler in patients with Parkinson’s disease. Ther Adv Chronic Dis. 2019 Jun 21; 10:2040622319857617. doi: 10.1177/2040622319857617. PMID: 31258882; PMCID: PMC6589987.

20. Grosset DG, Dhall R, Gurevich T, Kassubek J, Poewe WH, Rascol O, Rudzinska M, Cormier J, Sedkov A, Oh C. Inhaled levodopa in Parkinson’s disease patients with OFF periods: A randomized 12-month pulmonary safety study. Parkinsonism Relat Disord. 2020 Feb; 71:4–10. doi: 10.1016/j.parkreldis.2019.12.012. Epub 2019 Dec 23. PMID: 31927343.

21. Jost WH, Kulisevsky J, LeWitt PA. Inhaled levodopa for threatening impending OFF episodes in managing Parkinson’s disease. J Neural Transm (Vienna). 2023 Jun;130(6):821–826. doi: 10.1007/s00702-023-02636-3. Epub 2023 Apr 23. PMID: 37087697.

22. U R Anoop, Verma K: New technique and device for controlled and continuous drug delivery into the brain: a proof-of-concept study. BMJ Innovations 2021; 7:470–477.

23. Arunachalam JP, U R Anoop, Verma K, Rajendran R, Chidambaram S. SARS-CoV-2: The Road Less Traveled-From the Respiratory Mucosa to the Brain. ACS Omega. 2021 Mar 3;6(10):7068–7072. doi: 10.1021/acsomega.1c00030. PMID: 33748620; PMCID: PMC7970566.

24. Jayamuruga Pandian Arunachalam, Rahini Rajendran, Subbulakshmi Chidambaram, Srikanth Krishnagopal, S. Bhavani, V. Subha VeeranVeeravarmal, Harikrishnan Prasad, U.R Anoop, Kavita Verma. A novel maxillofacial technique to deliver drugs to the visual pathway, the optic nerve and the retina using the glymphatic system. bioRxiv 2022.03.23.485523; doi: https://doi.org/10.1101/2022.03.23.485523

25. Zlotnik Y, Balash Y, Korczyn AD, Giladi N, Gurevich T. Disorders of the oral cavity in Parkinson’s disease and parkinsonian syndromes. Parkinsons Dis. 2015; 2015:379482. doi: 10.1155/2015/379482. Epub 2015 Jan 15. PMID: 25685594; PMCID: PMC4312641.

